# The role of parenting on child socioemotional development: evidence from the 2015 Pelotas (Brazil) Birth Cohort Study

**DOI:** 10.1101/2025.04.22.25326200

**Authors:** Adriane Arteche, Otávio Amaral de Andrade Leão, Rafaela Costa Martins, Lynne Murray, Tiago N Munhoz, Roberta Salvador-Silva, Marlos Rodrigues Domingues, Joseph Murray

## Abstract

The influence of parenting on children’s socioemotional development at age 4 years was investigated using data from the 2015 Pelotas (Brazil) Birth Cohort. A total of 4010 children (50.6% male; mean age=45.5 months, SD=2.6; 72.4% White, 27.4% Black, 0.2% others) and their mothers (mean age=31.4 years, SD=6.6) took part in the study. Parenting was assessed via the Parental and Family Adjustment Scale and three observational tasks. Child mental health was assessed using the emotional symptoms, conduct problems, and hyperactivity/inattention subscales of the Strengths and Difficulties Questionnaire. Adjusted models showed that poor parental consistency, use of coercive parenting, and poor parent-child interactions were significantly associated with all three child outcomes.

---

Mental health disorders affect between 10% and 20% of all children and adolescents worldwide (Polanczyk et al., 2015), and previous research has shown that child emotional and behavioural problems have long lasting effects on social, educational, and later mental health outcomes (Belsky, 1984; Bowlby, 1969; Miller-Lewis et al., 2013; Sellers et al., 2019). This is of particular concern in low- and middle-income countries (Kieling et al., 2011) where healthcare support is scarce (Renwick et al., 2022) and exposure to chronic adversities is higher (Benjet, 2010).

A key environmental factor known to contribute to child mental health is parenting. Parenting is a broad dimension that encompasses patterns of attitudes and behaviors used to socialise the child as well as to encourage or reduce specific behaviors (Bornstein, 2013). Parenting and, in particular, mother–child interactions, operate as a context for children’s socio-emotional and cognitive development, directly affecting their emotion recognition and being a central mechanism on pathways to positive or dysfunctional outcomes (Phua, Kee & Meaney, 2020). Parenting is typically assessed in two broad dimensions: positive parenting, such as sensitivity (Shin et al., 2008), and negative or poor parenting, such as harsh behaviour (Runyan et al., 2010).

Sensitivity is the ability to identify, understand, and contingently respond to the child’s needs (Ainsworth, Blehar, Waters, & Wall, 1978). Sensitivity has been largely assessed via naturalistic or lab-filmed interactions between carer and child, with coding schemes including overall scores of the dyad’s interaction and specific behaviour expressed by carer, child, or dyad (Mesman, 2010). Regardless of the scoring system used, maternal sensitivity has been associated with better outcomes like secure attachment (Deans, 2020; O’Neill et al., 2021; Posada et al., 2016), good emotion regulation skills (Halligan, Cooper, Wheeler, Crosby, & Murray, 2013; Samdan et al., 2020), better academic achievement (Raby et al., 2015) and lower rates of emotional symptoms and behaviour problems (Sulik et al., 2015) – even in adverse environments (Murphy, Zhang, & Gatzke-Kopp, 2021). Specifically, maternal sensitivity when the child is young has been associated with lower rates of child externalising (Bernier et al., 2021) and internalising problems (Kok et al., 2013) during preschool years.

By contrast, harsh parenting refers to coercive acts and negative emotional expressions of parents towards their children and comprises both verbal and physical aggression (Hinnant et al., 2015) that may negatively affect child social and emotional development (Baydar & Akcinar, 2018; Berthelon, 2020; Chang et al, 2003; Rose et al., 2018). For instance, a meta-analysis using data from 971 studies found significant associations between harsh parenting and higher rates of child internalising and externalising symptoms, regardless of cultural differences in levels of acceptance of physical aggression (Pinquart, 2021).

Notably, several contextual factors may influence how parenting and, in particular, maternal nurturing, impact child mental health outcomes. Maternal sensitivity, for instance, may be particularly compromised by poverty, whereby parents invest all their efforts in attending to basic survival needs such as feeding, and lack skills to provide emotional support and responsiveness (Booth, Macdonald & Youssef, 2018). Neuhauser (2018), in an at-risk Swiss sample, revealed that risk factors such as lack of social support, low maternal education, and parental stress also contribute to lower levels of maternal sensitivity.

Finally, parenting (Bernard et al., 2018) and parental reports of child emotional and behavioural problems (Datta Gupta, Lausten, & Pozzoli, 2018) may be affected by maternal mental health, especially maternal depression. Although context is critical, most studies reporting the effects of parental sensitivity and harsh parenting on child mental health have been conducted in high-income countries. Evidence based on specific types of parenting and its impact on child development is sparse in Brazil and other low- and middle-income countries. Attempts to fill this gap have been carried out recently. Mazeyra et al (2022) analysed longitudinal data from the Young Lives project in Peru, conducted in a region where poverty rates range between 30% and 50%, and showed that early maternal sensitivity and play were both positively related to cognitive skills and socio-emotional competencies at ages 8 and 15 years. Results from the 2004 Pelotas Birth Cohort/Brazil revealed that harsh parenting assessed via the Conflict Tactic Scale at age six predicted emotional problems at age 11 (Bauer et al, 2021). Nevertheless, data on preschool children and observational data to measure parental practices (as well as parental self-reports) are lacking and may provide key evidence to inform early childhood interventions to support vulnerable families in adverse environments.

This study investigated the impact of positive (i.e., sensitivity) and negative (i.e., harsh) parenting on children’s socioemotional development at age 4 using data from the 2015 Pelotas Birth Cohort. Large studies of parenting hardly ever use observational measures, and, to our knowledge, this is the first large study using cohort data of a middle-income country to provide evidence about the associations between parenting and child socioemotional development using observational data and questionnaires.

## Methods

### Study Population and Participants

The 2015 Pelotas Birth Cohort is a longitudinal, population-based study conducted in Pelotas, a Southern Brazilian city with nearly 340,000 inhabitants. All women living in the urban area who gave birth in the city’s five maternity wards between January 1 and December 31, 2015, were invited to participate (around 99% of all births in Pelotas take place in these hospitals). In all, 4275 (98.7% of all live births of that year) children were enrolled in the cohort and have been followed ever since. The current study uses data collected in the perinatal and 4-year follow-ups, including 3997 children for whom outcome data were available and their primary caregiver, usually the child’s mother. Two approvals were obtained by the Research Ethics Committee of the Federal University of Pelotas (School of Physical Education #26746414.5.0000.5313 and Medical School #03837318.6.0000.5317) for perinatal and age four follow-ups. Caregivers provided written informed consent at each study follow-up. Further details of the 2015 Pelotas Cohort can be found elsewhere (Hallal et al., 2018; Murray, 2024).

## Measures

### Child Outcomes

*Strengths and Difficulties Questionnaire* (SDQ; Goodman, 1999; Goodman, Ford, Simons, Gatward & Meltzer, 2000): The SDQ is a questionnaire used to assess child emotional and behavioral difficulties that can be administered to the parents of 2- to 17-year-old children, previously validated in Brazil (Fleitich & Goodman, 2004; Fleitlich, Cortazar &, Goodman, 2000). The standard 25-item version is scored on a Likert-type scale ranging from 0 to 2 (0= not true; 1= somewhat true; 2= certainly true) and was completed by mothers in this study. For analyses, continuous scores and dichotomous categories of the emotional symptoms, conduct problems, and hyperactivity/inattention problems subscales were used. For categorical analyses, to establish clinical risk, the original (Goodman, 1999) three-band categories were applied as follows: emotional symptoms ≥5, conduct problems ≥3, and hyperactivity/inattention ≥6.

### Parenting

Parental and Family Adjustment Scale – PAFAS (Sanders, Morawska, Haslam, Filus & Fletcher, 2014; Santana, 2018): the 30-item original PAFAS consists of two domains [parenting practices (18 items) and family adjustment (12 items)]. The parenting practices domain encompasses four scales, including parental consistency (5 items), coercive parenting (5 items), positive encouragement (3 items), and parent-child relationship (5 items). This study used only the parenting practices domain, adopting a reduced version proposed by Martins et al. (2020) in which the final domain included 14 items (parental consistency = 3 items, coercive parenting = 4 items, positive encouragement = 2 items, and parent–child relationship = 5 items). The reduced version showed a good reliability coefficient (0.912). Each item is rated on a 4-point scale from ‘not at all’ (0) to ‘very much’ (3), and greater scores are associated with worse performance in all scales.

Book-sharing task (Cooper et al., 2014; Murray et al., 2016): this task aims to evaluate the mother’s sensitive responses and engagement in reciprocal exchanges with the child while sharing a book. Mother and child were filmed for approximately 5 minutes without examiner interference while looking at a picture book together. The book used was “A day in the park” — a book without text developed by the research staff to represent local activities and include content that may elicit dialogue of particular interest for the research project (e.g., children playing and conflict breaking out over toys in a sandpit). The coding was developed at the Winnicott Research Unit, University of Reading, and has been successfully applied in several previous studies (Cooper et al., 2014; Dowdall et al., 2021; Murray et al., 2016; Vally, Murray, Tomlinson & Cooper, 2015). The coding scheme assesses four dimensions: (A) child measures, (B) parent measures, (C) joint measures, and (D) specific book-sharing codes. Final scores are derived as either rating scales (usually 1-5) or event counts, with each dimension composed of specific sub-items. For the current study, parent measures of sensitivity and emotional tone were used – both scored in terms of overall behavior during the task. Sensitivity refers to parental awareness of the child’s direction of interest, behavioral cues, and appropriate and timely responsiveness. This dimension is coded on a 5-point scale: 1 (highly insensitive) to 5 (highly sensitive). Emotional tone refers to how happy and irritable the parent was during book-sharing and was also rated on a 5-point scale (1 = very unhappy to 5 = very happy).

Responsive Interactions (RI) task (Schneider, 2021): the RI task measures the overall construct of responsive interactions between carer and child, highlighting three interconnected skills of the caregiver: (i) communicative clarity (providing meaningful verbal/non-verbal inputs to the child and fostering of shared understanding of the goals of the task); (ii) mind reading (thinking about what the child knows and understands); and (iii) mutuality building (promoting reciprocity). The mother was instructed to sit and play with her child for five minutes in a semi-structured play activity that was video recorded. The mother and child were asked to construct a robot using play (Lego/Duplo type blocks) as per pictures shown. The mother and child should start with only the mom touching the Legos colored X and Y, and the child only touching the Legos colored Z and W. The mother was instructed to teach her child if the child had any doubts. For video coding, coders watched the 5-minute video recording of the activity only once, applying codes to each of the 11 items using a 5-point Likert scale, ranging from 1 (‘not at all true’) to 5 (‘very true’). Then, the mean result of the 11 items was calculated.

“Don’t Touch” – prohibition – task (DTT; Cooper, 2017; Kochanska & Aksan, 1995): the DTT aims to assess harsh parenting and child frustration. Mother and child are presented with boxes full of interesting toys. The examiner removes some toys from these boxes and puts them in front of the child, instructing that the child is not allowed to touch them (nor the mother). After approximately 2 minutes and 30 seconds, the child is allowed to play with only one toy. The task ends after a further 1 minute. Scoring comprises one dimension of child behavior and four dimensions of carer behavior. For the current study, an overall score of coercive parenting was used. This was composed by the sum scores of coercive, harsh, control/discipline verbal (event count of verbal threat/coercive control for the whole period of the task) and coercive, harsh, control/discipline physical (scores range 0-3; degree of physical coercive control, rated for each 20 second time block with a score of 3 meaning that the carer shows at least one episode of clear directed aggression). The task had been previously adapted and used in the PIÁ trial in the same population as the current study (Murray et al., 2019; Murray et al., 2024).

### Covariates

Information on child sex (male, female) was obtained from the perinatal assessment. Maternal age (<20, 20-34, >25), maternal schooling in years (0-4, 5-8, 9-12, >12), current maternal status of work (not working, working), father presence in the house (no father at home, biological father at home, social father at home) and maternal relationship status (no partner, with partner) was obtained at 4-year data collection via a sociodemographic questionnaire.

Neighbourhood violence was measured using maternal reports on the frequency (0 = never to 3 = often) of the following types of violence in the neighbourhood in the last 6 months: fights with weapons, fights between gangs, robbery, and sexual violence.

Scores were summed, and neighbourhood violence was classified in three categories (0-2, 3-7, and ≥8). Family income was assessed as a continuous variable and obtained by summing the monthly income of all household members and then dividing the total into quintiles. Maternal alcohol drinking and drug use were assessed at the 4-year follow-up using the Alcohol, Smoking and Substance Involvement Screening Test (ASSIST; WHO ASSIST Working Group, 2002; Humeniuk et al., 2018). For the Brazilian validation of the ASSIST questionnaire, see Henrique et al. (2004). Drug use was characterised as the use of any illicit substance in the three months before the interview, and alcohol use was defined as daily drinking of any alcoholic beverage.

Maternal postnatal depression was assessed at 3 months postpartum using the Edinburgh Postnatal Depression Scale (EPDS), which was validated in a Brazilian population (Santos et al., 2007). Mothers with scores ≥ 13 were defined as depressed. Mother-partner relationship conflict was also assessed at the 3-month follow-up using two Likert-type questions about partners’ criticism of each other (Hooley, Orley & Teasdale, 1986), ranging from 1 to 10 (low to high criticism). Questions assessed how critical the mother considered her partner and how critical she considered herself to be of her partner. Following Buffarini et al. (2021), we first categorised each individual score as reflecting low (1-3), medium (4-6), or high (7-10) criticism, and then combined the two into a relationship criticism score coded as follows: low criticism (low-low or low-medium); medium criticism (medium-medium, or low-high), or high criticism (medium-high or high-high).

### Procedures

The 4-year follow-up of the 2015 Pelotas Birth Cohort occurred between February and August 2019. Data collection took place at the University research clinic, lasted about 3 hours, and was conducted by a team of trained research assistants. All observational tasks were filmed. Coding was carried out by a team of graduate psychologists or undergraduate research assistants as follows: four coders for the Booksharing task, three coders for the Don’t touch task, and three coders for the Responsive Interactions task. For the Booksharing and the Don’t touch coding, work was done by trained undergraduate psychologists under the supervision of a senior psychologist. Training involved a presentation of the measures with examples and group coding of videos. Coders then received blocks of 10 videos to code, which the senior psychologist reviewed. Disagreements were discussed until an agreement of 80% or more was achieved. After initial training, the main coding proceeded with coders leaving notes about any doubts discussed in the regular weekly supervision meetings. Additionally, the senior psychologist coded a random sample of 10% of the videos to ensure quality control and calculate reliability. Inter-coder kappa coefficients were 0.97 and 0.99 for Booksharing Sensitivity and Don’t Touch Coercion, respectively. For the Responsive Interaction task, training was conducted via the online platform and supervised by the instrument developer of the Brazilian version. A gold standard coder who reached 100% agreement in the online training carried out regular checks of the coding procedures and quality control. The intraclass correlation coefficient for a random sample of 10% of the videos was 0.62.

### Statistical Analysis

Descriptive analyses were used to summarise the sample characteristics, using absolute and relative frequencies for categorical variables and mean and standard deviation for numerical variables. To improve the interpretability of the results, descriptive analyses for all variables were run, including only children who had valid data for the study outcomes. The associations between parental exposures (PAFAS and observational parenting tasks) and child outcomes (SDQ) were examined using Negative Binomial Regression Models (NBRM), given the count characteristic of the outcomes. An Incident Rate Ratio (IRR) effect size was calculated by exponentiation of the NBRM regression coefficients. These represent the relative difference in the child outcome score for a unit increase in the parenting exposure variable. Both crude and adjusted NBRM models were run. The adjusted model included all parenting exposures and socioeconomic confounders (maternal scholarship, family income, maternal postnatal depression, and mother-partner relationship) with p<0.20. To improve the interpretability of the adjusted model, Pseudo R², p-value, Akaike’s Information Criteria (AIC), and Bayesian Information Criteria (BIC) are presented. Additional sensitivity analyses were performed using adjusted logistic models, based on risk variables of the outcomes. For all models, 95% confidence intervals were presented. All analyses were conducted using STATA 16.0, with statistical significance set at 5%.

## RESULTS

Children’s (mean age 45.5 months, SD=2.6; 72,4% White, 27.4% Black, 0.2% other ethnicity) and mothers’ characteristics (mean age 31.4 years, SD=6.6) are presented in Table 1.

**Table 1.**
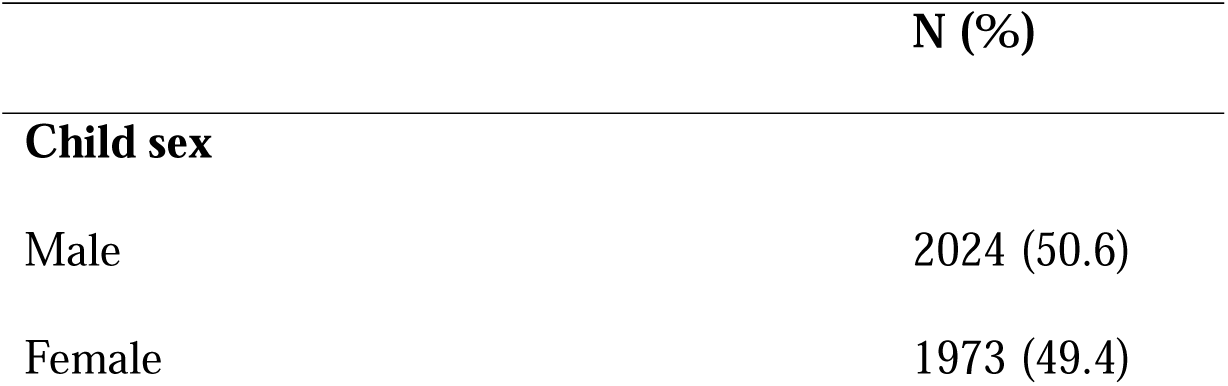
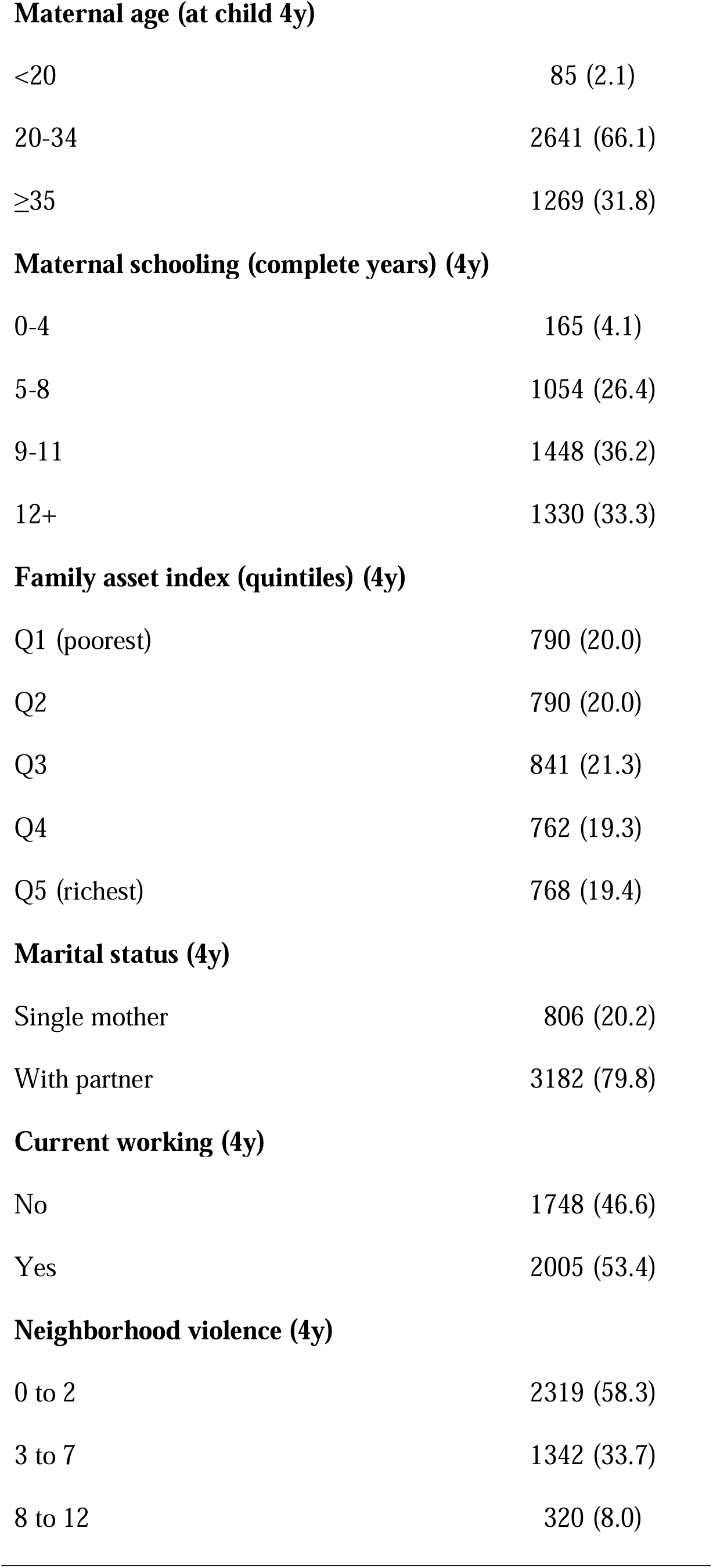
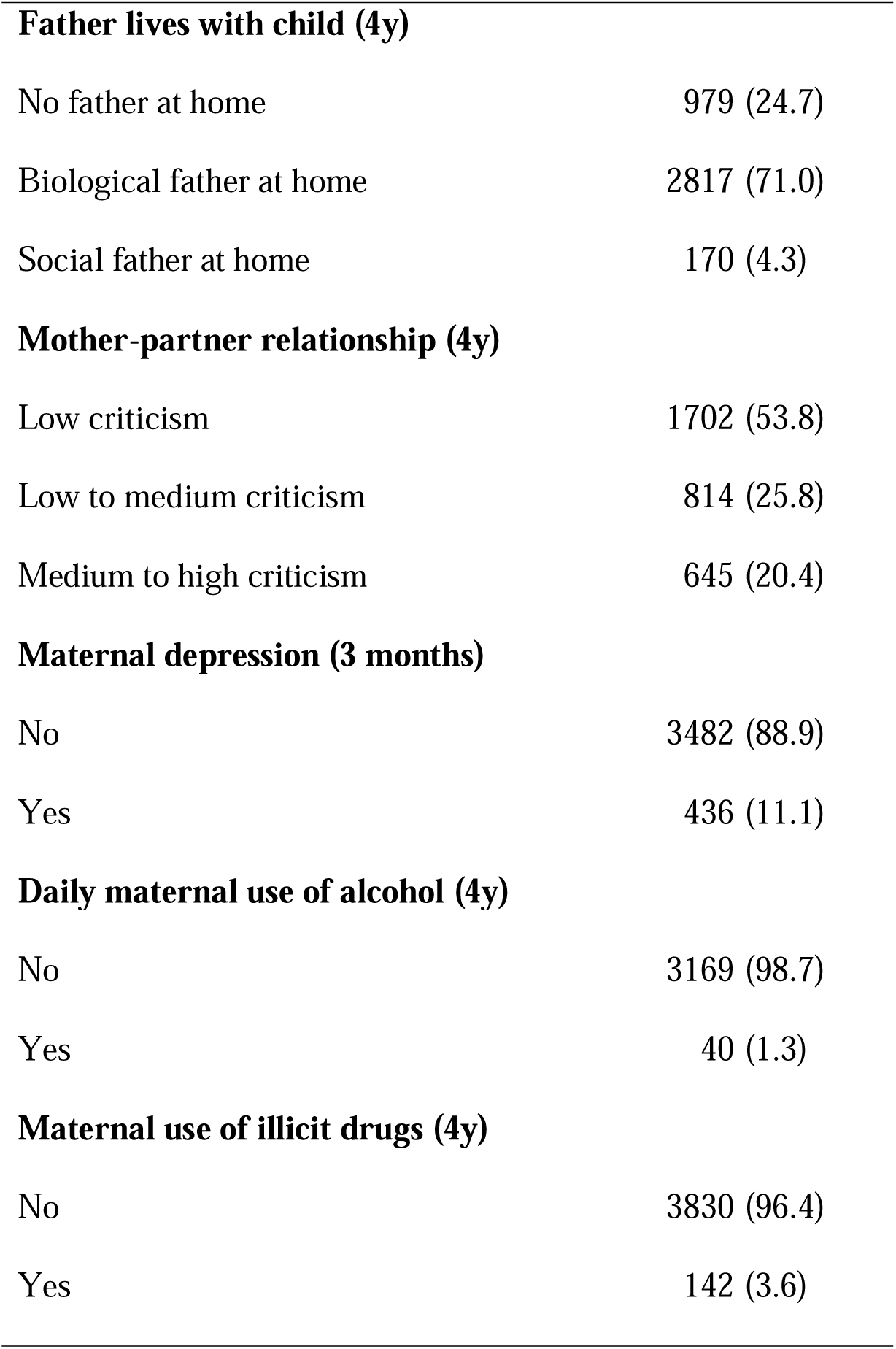
Descriptive sample characteristics (N=3997).

|  | N (%) |
| --- | --- |
| **Child sex** |  |
| Male | 2024 (50.6) |
| Female | 1973 (49.4) |
| **Maternal age (at child 4y)** |  |
| <20 | 85 (2.1) |
| 20-34 | 2641 (66.1) |
| ≥35 | 1269 (31.8) |
| **Maternal schooling (complete years) (4y)** |  |
| 0-4 | 165 (4.1) |
| 5-8 | 1054 (26.4) |
| 9-11 | 1448 (36.2) |
| 12+ | 1330 (33.3) |
| **Family asset index (quintiles) (4y)** |  |
| Q1 (poorest) | 790 (20.0) |
| Q2 | 790 (20.0) |
| Q3 | 841 (21.3) |
| Q4 | 762 (19.3) |
| Q5 (richest) | 768 (19.4) |
| **Marital status (4y)** |  |
| Single mother | 806 (20.2) |
| With partner | 3182 (79.8) |
| **Current working (4y)** |  |
| No | 1748 (46.6) |
| Yes | 2005 (53.4) |
| **Neighborhood violence (4y)** |  |
| 0 to 2 | 2319 (58.3) |
| 3 to 7 | 1342 (33.7) |
| 8 to 12 | 320 (8.0) |
| **Father lives with child (4y)** |  |
| No father at home | 979 (24.7) |
| Biological father at home | 2817 (71.0) |
| Social father at home | 170 (4.3) |
| **Mother-partner relationship (4y)** |  |
| Low criticism | 1702 (53.8) |
| Low to medium criticism | 814 (25.8) |
| Medium to high criticism | 645 (20.4) |
| **Maternal depression (3 months)** |  |
| No | 3482 (88.9) |
| Yes | 436 (11.1) |
| **Daily maternal use of alcohol (4y)** |  |
| No | 3169 (98.7) |
| Yes | 40 (1.3) |
| **Maternal use of illicit drugs (4y)** |  |
| No | 3830 (96.4) |
| Yes | 142 (3.6) |

Table 2 presents descriptive statistics for child outcomes and parenting measures. Continuous child outcomes scores were inspected, and the highest means were observed for hyperactivity, followed by conduct and emotional subscale scores. Descriptive results for the parenting dimensions assessed by the PAFAS suggested low levels of poor parenting. Parenting dimensions assessed via filmed tasks revealed mean scores similar to each task’s midpoint on the scale (e.g., mean 2.53 for the Responsive Interactions task with a range of 0-5).

**Table 2.** Exposures and outcomes descriptives for 2015 Pelotas Birth Cohort participants at age 4.

|  | Mean (SD) | Range |
| --- | --- | --- |
| **Parenting** |  |  |
| *PAFAS* |  |  |
| Consistency<sup>+</sup> | 2.00 (1.74) | 0 – 9 |
| Positive Encouragement<sup>+</sup> | 0.79 (0.97) | 0 – 6 |
| Parent-child relationship<sup>+</sup> | 1.30 (1.85) | 0 – 12 |
| Coercive Parenting | 3.59 (2.08) | 0 - 12 |
| *Filmed tasks* |  |  |
| Responsive Interactions | 2.53 (0.83) | 1 – 4.7 |
| Emotional tone<sup>+</sup> | 2.99 (0.70) | 0 – 5 |
| Sensitivity | 3.62 (0.85) | 1 – 5 |
| Coercive Total | 0.58(0.75) | 0- 3 |
| **Child Outcomes** |  |  |
|  | Mean (SD) | Range |
| Emotional problems (0-10) | 2.38 (1.98) | 0 – 10 |
| Conduct problems (0-10) | 2.49 (2.03) | 0 – 10 |
| Hyperactivity (0-10) | 4.29 (3.06) | 0 – 10 |
|  | **n** | **%** |
| Emotional problems |  |  |
| Yes | 586 | 14.7 |
| No | 3412 | 85.3 |
| Conduct problems |  |  |
| Yes | 1216 | 30.4 |
| No | 2782 | 69.6 |
| Hyperactivity |  |  |
| Yes | 1038 | 26.0 |
| No | 2959 | 74.0 |

<sup>†</sup>Higher scores in Consistency, Encouragement, and Parent-Child Relationship depict a worse outcome. Higher scores in emotional tone depict a better outcome.

Table 3 shows the unadjusted predictive model of child emotional, conduct, and hyperactivity problems considering each parenting measure as an exposure. Parenting dimensions were significantly associated with child outcomes, except for coercive parenting assessed through the Dońt Touch task, which was not significantly associated with emotional problems.

**Table 3.**
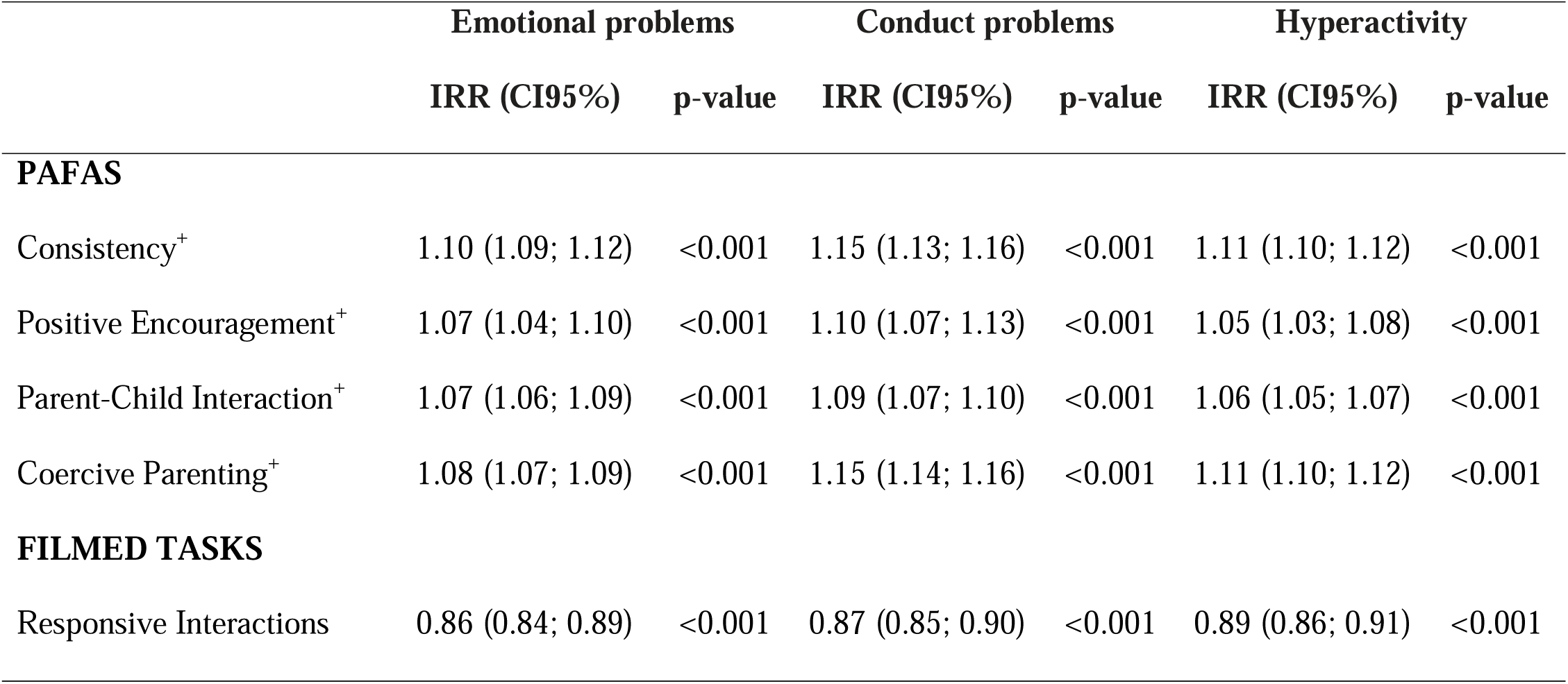
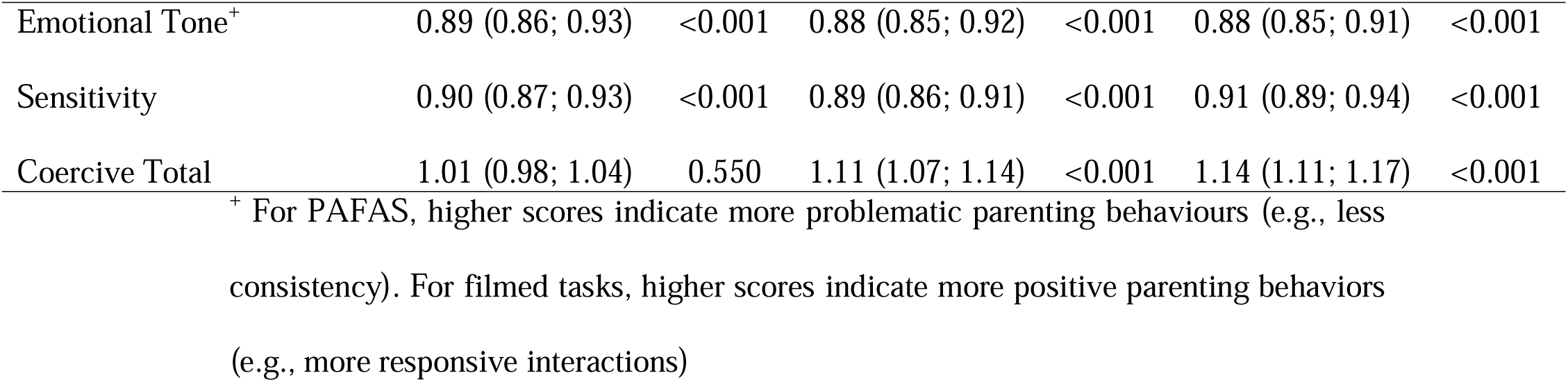
Unadjusted associations between parenting and child mental health using negative binomial regression models.

|  | Emotional problems |  | Conduct problems |  | Hyperactivity |  |
| --- | --- | --- | --- | --- | --- | --- |
|  | IRR (CI95%) | p-value | IRR (CI95%) | p-value | IRR (CI95%) | p-value |
| **PAFAS** |  |  |  |  |  |  |
| Consistency<sup>†</sup> | 1.10 (1.09; 1.12) | <0.001 | 1.15 (1.13; 1.16) | <0.001 | 1.11 (1.10; 1.12) | <0.001 |
| Positive Encouragement<sup>†</sup> | 1.07 (1.04; 1.10) | <0.001 | 1.10 (1.07; 1.13) | <0.001 | 1.05 (1.03; 1.08) | <0.001 |
| Parent-Child Interaction<sup>†</sup> | 1.07 (1.06; 1.09) | <0.001 | 1.09 (1.07; 1.10) | <0.001 | 1.06 (1.05; 1.07) | <0.001 |
| Coercive Parenting<sup>†</sup> | 1.08 (1.07; 1.09) | <0.001 | 1.15 (1.14; 1.16) | <0.001 | 1.11 (1.10; 1.12) | <0.001 |
| **FILMED TASKS** |  |  |  |  |  |  |
| Responsive Interactions | 0.86 (0.84; 0.89) | <0.001 | 0.87 (0.85; 0.90) | <0.001 | 0.89 (0.86; 0.91) | <0.001 |
| Emotional Tone<sup>+</sup> | 0.89 (0.86; 0.93) | <0.001 | 0.88 (0.85; 0.92) | <0.001 | 0.88 (0.85; 0.91) | <0.001 |
| Sensitivity | 0.90 (0.87; 0.93) | <0.001 | 0.89 (0.86; 0.91) | <0.001 | 0.91 (0.89; 0.94) | <0.001 |
| Coercive Total | 1.01 (0.98; 1.04) | 0.550 | 1.11 (1.07; 1.14) | <0.001 | 1.14 (1.11; 1.17) | <0.001 |

<sup>+</sup> For PAFAS, higher scores indicate more problematic parenting behaviours (e.g., less consistency). For filmed tasks, higher scores indicate more positive parenting behaviors (e.g., more responsive interactions)

As seen in Table 4, the models adjusting for all other risk factors retained parental poor consistency, the use of coercive parenting and poor parent-child interactions, as assessed via the PAFAS, as significant predictors of all three child outcomes — with relative increases in child problem scores ranging from 1.03 to1.11 for each unit increase in parenting exposures. Considering the PAFAS exposures, the strongest association was between coercive parenting and conduct problems, while in observational tasks the largest effect size was between coercive parenting (measured by the Don’t Touch task) and hyperactivity.

**Table 4.** Adjusted associations between parenting and child mental health using negative binomial regression models.

|  | Emotional problems |  | Conduct problems |  | Hyperactivity/Inattention |  |
| --- | --- | --- | --- | --- | --- | --- |
|  | IRR (CI95%) | p-value | IRR (CI95%) | p-value | IRR (CI95%) | p-value |
| **PAFAS** |  |  |  |  |  |  |
| Consistency<sup>+</sup> | 1.04 (1.02; 1.05) | <0.001 | 1.08 (1.07; 1.10) | <0.001 | 1.06 (1.04; 1.07) | <0.001 |
| Positive encouragement<sup>+</sup> | 0.97 (0.94; 1.01) | 0.17 | 1.02 (0.99; 1.06) | 0.14 | 0.98 (0.95; 1.02) | 0.34 |
| Parent-child relationship<sup>+</sup> | 1.04 (1.02; 1.06) | <0.001 | 1.04 (1.02; 1.06) | <0.001 | 1.03 (1.01; 1.04) | 0.001 |
| Coercive parenting<sup>+</sup> | 1.05 (1.03; 1.06) | <0.001 | 1.11 (1.09; 1.12) | <0.001 | 1.08 (1.07; 1.10) | <0.001 |
| **FILMED TASKS** |  |  |  |  |  |  |
| Responsive interactions | 0.98 (0.94; 1.02) | 0.31 | 0.99 (0.96; 1.03) | 0.79 | 0.99 (0.96; 1.03) | 0.77 |
| Emotional tone | 1.00 (0.95; 1.04) | 0.91 | 0.98 (0.93; 1.02) | 0.35 | 0.94 (0.90; 0.98) | 0.007 |
| Sensitivity | 0.99 (0.95; 1.03) | 0.64 | 0.99 (0.95; 1.03) | 0.58 | 1.02 (0.98; 1.05) | 0.35 |
| Coercive total | 0.98 (0.94;1.02) | 0.31 | 1.06 (1.02; 1.09) | 0.002 | 1.12 (1.08; 1.16) | <0.001 |
| **Model values** |  |  |  |  |  |  |
| p-value | <0.001 |  | <0.001 |  | <0.001 |  |
| Pseudo R<sup>2</sup> | 0.03 |  | 0.06 |  | 0.03 |  |
| AIC | 11181.96 |  | 11009.34 |  | 14063.08 |  |
| BIC | 11265.79 |  | 11093.17 |  | 14146.90 |  |

\*Adjusted for all parenting exposures and socioeconomic variables with p<0.20: maternal scholarship, family income, maternal postnatal depression, and mother-partner relationship. <sup>+</sup>For PAFAS, higher scores indicate more problematic parenting

Also in Table 4, coercive parenting assessed via observation significantly increased mean scores of child conduct problems (IRR = 1.06; 95%CI: 1.02 - 1.09) and hyperactivity problems (IRR = 1.12; 95%CI: 1.08 - 1.16), whereas higher scores in parental positive emotional tone significantly decreased mean levels of hyperactivity problems (IRR = 0.94; 95%CI: 0.90 - 0.98). Interestingly, parental use of coercive practices seems to play an important role regardless of whether it is assessed via the PAFAS or the observational task.

As expected, the three comprehensive models, including all parenting measures as predictors of each child outcome, were significant, with better AIC indices for the conduct problems model. Analyses revealed that, using a Pseudo R², 6% of the variance in child conduct problem scores and 3% of the variance of child emotional and hyperactivity problem scores were explained by the models.

When dimensions of parenting were explored as predictors of risk for a dichotomous status of child emotional, conduct, and hyperactivity problems, the effects of parental use of coercive practices stand out for all three child outcomes, even when adjusting for other potential risk factors (see Figure 1). The chance of a child being classified as at risk for emotional problems increased by 15% for each additional point of parental coercive practices’ subscore of the PAFAS, by 12% for each point increase in the parent-child relationship subscale score, and by 10% for each increase in the (in)consistency subscale. Increases in the coercive subscale are associated with a 29% greater chance of the child having a conduct problem and higher scores in the (in)consistency subscale are associated with a 22% increase in chances of a child having a conduct problem. Finally, children of parents with higher coercive parenting scores in the Dońt Touch observational task have 36% more chance of being classified as at risk for conduct problems; 29% greater risk was associated with higher scores on the PAFAS coercive subscale, and 22% greater risk with the PAFAS (In)consistency subscale.

**Figure 1.**
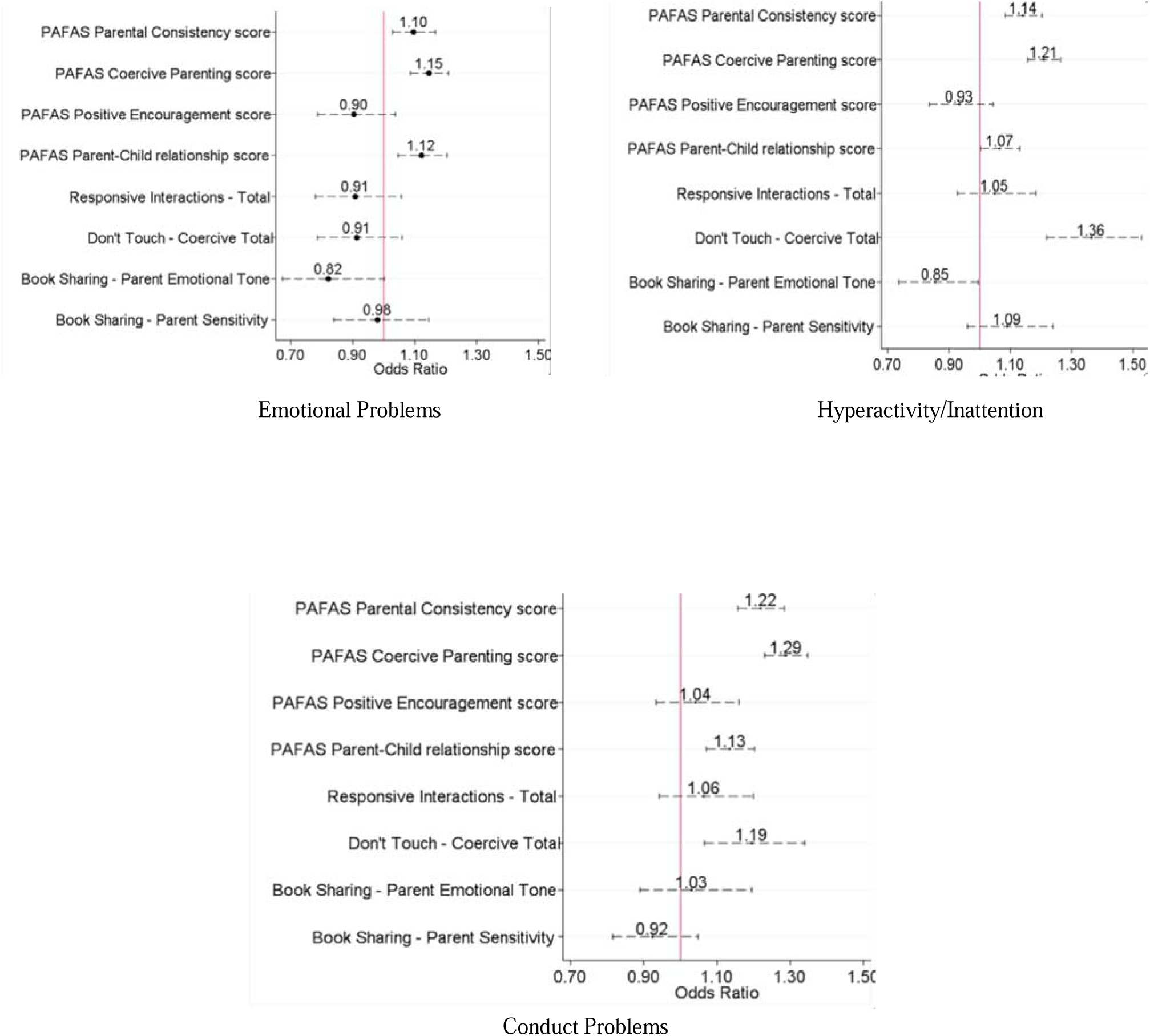
Adjusted associations of parental characteristics and risk of emotional problems, conduct problems, and hyperactivity using Logistic Regression. *Adjusted for all parenting exposures and socioeconomic variables with p<0.20: maternal scholarship, family income, maternal postnatal depression, and mother-partner relationship.

## Discussion

This study aimed to shed light on the associations between parenting and child mental health (i.e., emotional, conduct, and hyperactivity problems). We assessed positive and negative parenting dimensions using both questionnaire and observational data. Additionally, we used a large sample from South Brazil, providing unique data on parental dimensions and child outcomes in a developing country. Accordingly, this study fills a gap in research in low- and middle-income countries regarding parenting and child mental health in early childhood. Still, it also addresses a worldwide lack of evidence from population-based large studies on this theme, especially using observational data.

Using the Strengths and Difficulties Questionnaire, levels of child emotional, conduct, and hyperactivity problems found in the current study confirmed previous literature (Kieling et al., 2011), suggesting high rates of mental health problems even in preschool children. Notably, using Goodman’s (1999) clinical risk threshold, close to 15% of our sample scored above the cut-off point for emotional problems, and rates of conduct and hyperactivity problems were up to 30%. High rates of conduct problems in Brazilian samples have been previously discussed in Murray et al. (2013) as related to various factors, including cultural bias in parental reports and exposure to environmental violence. Risk factors associated with young people’s socioemotional difficulties include factors such as parental education, family income, maternal postnatal depression, and mother-partner relationship (Nolan & Smyth, 2021). In our study, when those were considered, parenting remained significantly associated with child emotional, conduct, and hyperactivity problems. This finding strengthens the key role of parenting even in a low- and middle-income country.

In particular, parenting dimensions retained in the model were those related to negative or harsh parenting. Poor parental consistency, the use of coercive parenting, and poor parent-child relationship assessed by the PAFAS and coercive parenting assessed via the Don’t Touch task were important predictors of child socioemotional development. Previous studies have shown that sociodemographic risk factors such as family income and maternal mental health may compromise maternal ability to positively provide emotional support and contingently identify child’s needs (Bernard et al., 2018; Booth, Macdonald & Youssef, 2018; Neuhauser, 2018). In our study, we showed that even when those factors are controlled for, the use of coercive or harsh parenting practices still plays an important role in children’s socioemotional development.

Additionally, negative parenting practices seem to have an important and broad effect on child socioemotional development. In our study, parental use of coercive practices was a key predictor of child socioemotional problems, regardless of whether it was assessed via questionnaire or filmed observation. The concerning use of coercive practices by parents has been documented in the Pinquart (2021) review, and our study expands on those findings, suggesting that those effects are independent of other risk factors and levels of positive parenting behaviours. This finding may be of particular importance for the development of intervention programs in Brazil given that previous research in Brazil has highlighted widely spread cultural beliefs that hitting a child or using harsh discipline is acceptable if parents want to teach something to the child (Medeiros et al., 2021; Murray et al., 2019).

A considerable proportion of mothers in our sample reported repeatedly and consistently using parenting practices that involve shouting, arguing, trying to make the child feel bad, and/or spanking or smacking. Even though mothers were aware of being filmed and observed during the observational tasks, the use of coercive practices still lasted. Across the globe, harsh parenting is associated with negative outcomes, but cultural moderators and nonlinear associations may operate in specific countries (Pinquart, 2021). In Brazil (Coll et al., 2023), the intergenerational problem of domestic violence is potentially mediated by the use of harsh parenting, which leads to higher rates of conduct problems during childhood, which in turn are themselves associated with greater rates of violence, including domestic violence in adulthood. Therefore, potential impacts of the observed use of harsh parenting may go beyond the current assessed participants and extend to future generations.

When the specific dimensions of parenting effects were inspected it is worth noting that in the unadjusted model both questionnaires and observational tasks were significant; however, in the adjusted models, most significant measures came from the PAFAS questionnaire. This may be related to the more general aspect of parenting assessed by the PAFAS, as opposed to a short time-slice of parenting behaviour observed in the tasks. Additionally, it could be that the PAFAS captures a broader picture of parental behaviour, whereas the observational tasks offer a more subtle version of parenting due to the presence of the observer and the specificity of the task itself. This is supported by the fact that coercive parenting assessed via the Dońt Touch observational task - which indeed gives more scope for an explicit and broader harsh parental behaviour remained a significant predictor of conduct and hyperactivity problems even in adjusted analyses. These findings also support the use of various strategies to assess parental practices in future investigations. This approach enabled us to provide a broader picture of specific effects of each parenting dimension.

Measures used in this study include questionnaires and observational tasks that have different ranges and hence unit changes have different applied implications; nevertheless, it is of note that parental consistency and parental use of coercive parenting were the strongest predictors of all three child outcomes: for every unit of increase, the child emotional, conduct and hyperactivity problems rate increased by 8-15%. Finally, when the different child socioemotional outcomes are inspected, it is notable that parenting had its strongest effects on behavioural difficulties, such as conduct problems and hyperactivity, with the best AIC indices being observed for the conduct problems model. Rates of conduct and hyperactivity problems were also twice as high as those of emotional problems. This may be explained by the child’s age and the use of a parental report measure. The onset of emotional problems in children is more frequent at school age (Ogundele, 2018). In contrast, behavioural difficulties are usually reported earlier in preschool children (Bagner, 2012). Bias and low child-parent agreement in parental reports of child difficulties have also been previously identified, with a tendency for parents to report more behavioural than emotional difficulties (van der Meer, Dixon, & Rose, 2008). Conversely, it is also plausible that rates of behavioural problems are indeed particularly associated with the observed high rates of coercive practices.

Our study used data from a city located in South Brazil and may not account for the cultural diversity of the country. Additionally, we used parental reports and not a clinical interview conducted by an independent clinician to establish rates of socioemotional problems. Also, our observational tasks were part of a set of assessments, and children and mothers could be tired in some of the tasks. Finally, one of the major limitations is the cross-sectional character of the analyses, which limits our interpretation of the findings. Despite those limitations, we provide unique data on parenting and child mental health with both questionnaires and observational measures from a large population study which has the potential to guide future interventions.

## CONCLUSION

Understanding the links between parenting and offspring emotional and behavioural difficulties and providing evidence of the particular dimensions of parenting that are most strongly associated with child mental health are key requirements for designing psychoeducational and clinical interventions involving parents. Supporting parents’ interactions with their offspring represents a unique opportunity to potentially improve mental health, not only in today’s families but also in generations to come. The findings presented in the current study show that parenting is a key mechanism linked to child mental health. Notably, the parenting dimension that was most importantly related to children’s conduct, hyperactivity, and emotional problems was coercive parenting. This finding indicates that interventions should focus on training parents on how to manage interactions with their children in more constructive ways, particularly in contexts where coercive practices are commonly adopted.

## Data Availability

All data produced in the present study are available upon reasonable request to the authors

## Acknowledgements

This article is based on data from the study "Pelotas Birth Cohort, 2015" conducted by Postgraduate Program in Epidemiology at *Universidade Federal de Pelotas*, with the collaboration of the Brazilian Public Health Association (ABRASCO). The first phases of the 2015 Pelotas (Brazil) Birth Cohort was funded by the Wellcome Trust (095582). Funding for specific follow-up visits was also received from the *Conselho Nacional de Desenvolvimento Científico e Tecnológico* (CNPq), *Fundação de Amparo a Pesquisa do Estado do Rio Grande do Sul* (FAPERGS), and Children’s Pastorate sponsored follow-up at twenty-four months; and FAPERGS–PPSUS, the Wellcome Trust (10735_A_18_Z), and the Bernard van Leer Foundation (BRA-2018-178) for the 4-year follow-up. At the 4-year follow-up, the 2015 cohort was also funded by the Department of Science and Technology (DECIT/Brazilian Ministry of Health). The data and materials necessary to reproduce the analyses presented here are not publicly accessible; data from the 2015 Pelotas Birth Cohort may be accessed by contacting the lead authors. The analytic code necessary to reproduce the analyses presented in this paper is accessible by asking the authors. The analyses presented here were not preregistered. This research was funded in whole, or in part, by the Wellcome Trust [210735_A_18_Z]. For open access, the author has applied a CC BY public copyright licence to any Author Accepted Manuscript version arising from this submission.

## Conflict of interest

None

## Notes

### Competing Interest Statement

The authors have declared no competing interest.

### Funding Statement

Wellcome Trust 095582, FAPERGS/PPSUS, Wellcome Trust 10735_A_18_Z, Bernard van Leer Foundation BRA-2018-178

### Author Declarations

Ethics Committee of UFPel gave ethical approval for this work (School of Physical Education #26746414.5.0000.5313 and Medical School #03837318.6.0000.5317)

